# Semen Analysis Accuracy and Consequences on Therapeutic Decisions

**DOI:** 10.1101/2025.05.08.25327189

**Authors:** Javier Barranco Garcia, Anna Kuzmina, Austin Lee, Jeyla Sadikova, Mahmoud Elhusseiny Mostafa, Run Zhang, Fabien Murisier, Véronique Vallet Conde, Loup Cordey, Michel Bielecki

## Abstract

Male-factor infertility accounts for nearly half of all infertility cases worldwide, yet conventional semen analysis methods—both manual and computer-assisted—can yield inconsistent results. Such inaccuracy can lead to misleading diagnosis, unnecessary interventions (e.g., IVF/ICSI), and potentially increased healthcare costs. Here, we present a novel imaging system, LuceDX, which uses a 13-fold expanded field of view (FOV) to overcome the statistical and technical limitations of standard Computer Assisted Semen Analysis (CASA) tools. By capturing a substantially larger sample area, LuceDX mitigates the non-uniform sperm distribution and clustering effects that compromise accuracy in smaller FOV methods. Pilot data indicate that this expanded-FOV platform improves measurement precision by a factor of 3.6 relative to conventional techniques, aligning with WHO guidelines while reducing the need for multiple fields per sample. This improvement is particularly advantageous in oligospermic men and post-vasectomy assessments, where accurate detection of very low sperm counts is critical in clinical decision making, especially considering real-world settings where guidelines may not be consistently applied. Our discussion highlights the patient and physician benefits of increasing accuracy and reliability toward an even better fertility treatment efficacy and underscores the importance of larger, multi-center trials to validate these findings. Taken together, the evidence suggests that LuceDX’s integrated, AI-enabled advanced image analysis offers a promising step forward in male infertility diagnostics, improving reliability, reducing costs, and potentially accelerating the optimal management of infertile couples.

## Introduction

Infertility represents a significant public health concern, with an estimated 48.5 million couples worldwide experiencing conception difficulties^1^ and in Europe alone, over 800,000 ART cycles starting yearly^2^. Male factors contribute to approximately 40–50% of these cases^3^. Despite this prevalence, the diagnostic infrastructure for male infertility remains limited, often leading to delayed evaluation and intervention.

Manual semen analysis, as outlined by WHO guidelines, is the standard first-line test for male infertility^4^. Even with extensive training, subjective differences, and intra-/inter-observer variability can be high, leading to inconsistent sperm counts, motility, and morphology assessments^4^. In fact, studies have documented large inter-laboratory variation in results – with reported coefficients of variation ranging from ∼23% up to 73% for sperm concentration measurements and similarly high variability in motility and morphology evaluations^5,6^. Such error rates directly threaten the accuracy of results and can impact clinical decision-making ^4^.

Computer-Assisted Semen Analysis (CASA) systems were introduced to reduce operator subjectivity and standardize measurements. These automated systems use imaging software to analyze semen parameters, allowing faster processing and improved consistency compared to manual analyses^4^. While CASA systems do improve reproducibility and throughput, studies show only marginal accuracy gains over manual analysis.^4,7,8^ Notably, CASA results can diverge from manual counts, especially in samples with very low or very high sperm counts, or abnormal motility. For example, one systematic review found strong correlations between manual and CASA measurements in normospermic samples but significantly poorer agreement in cases of moderate or severe oligozoospermia^4^. In other words, CASA may still produce highly variable results for abnormal samples, sometimes requiring frequent recalibration and technical oversight.^9^ Concerns remain about the validity and reliability of CASA versus the recommended manual method, particularly in low concentration specimens^4^.

However, one major limitation of both manual and CASA methods is linked to the limited volume of a sample that can be assessed using conventional microscopy techniques. Low concentration samples require the analysis of a substantial sample volume, which translates into analysing a large number of fields of view (FOVs). In practice, analyzing the additional sample volume required for low-concentration specimens is often skipped due to the time and effort involved. This selective examination biases results, yielding artificially high accuracy in normal samples while compromising the reliability of measurements in pathological cases, which are more prevalent in clinical practice.

Overall, both traditional manual analysis and current CASA techniques exhibit measurement uncertainty and error that can limit diagnostic reliability.

### Clinical Consequences of Inaccurate Semen Analysis

Intra-individual variations in semen parameters have been described in several publications^10–12^. These variations could be seasonal or linked to not always identified external causes such as fever or medications. Inaccurate semen analysis might also play a role in these findings. It is recommended, in case of anomalies, to perform a second semen analysis three months later given that the time required for the sperm to form is around 74 days. A study showed that in one quarter of the cases, the second exam did not confirm the initial diagnosis.^13^ This could be particularly misleading when the first semen analysis is normal and thus not repeated.

When semen analysis results are inaccurate, the risk of misdiagnosis of a couple’s infertility etiology rises, leading to improper management. This is particularly crucial as semen analysis – while technically speaking is not a functional test of fertility and rather a reflection of spermatogenesis and the surrounding physiologic milieu – but at present is the only evaluation method for assessment of male infertility available to clinicians.. Key consequences of semen analysis imprecisions include:

- **Unnecessary Invasive Procedures:** Erroneous abnormal results in the male may prompt invasive interventions that aren’t actually needed. For instance, a falsely poor semen analysis might push a couple toward costly assisted reproduction (e.g. IVF/ICSI) or lead to surgeries like varicocelectomy based on incorrect data. Conversely, missing a male factor problem can subject the female partner to invasive fertility treatments unnecessarily^14^. In short, inaccurate semen analysis can misdirect couples to invasive ART or surgeries that could have been avoided.
- **Suboptimal or Delayed Treatments:** A wrong semen diagnosis may focus treatment on the wrong cause or delay the appropriate intervention. For example, a borderline abnormal result that isn’t confirmed can lead physicians to pursue additional male or female diagnostic tests that aren’t needed, wasting time. A recent clinical analysis underscored that not confirming an initial semen analysis with a second test can result in unnecessary examinations and treatment delays^15^. Likewise, clinicians must recognize the “intrinsic uncertainties” of a single semen test; if they don’t, they might treat based on a flawed result and postpone the correct therapy for the couple ^9^. This can prolong the period a couple remains infertile. Thus, poor accuracy in semen analysis may cause misdirected therapies or slow down the use of effective treatments, directly impacting a couple’s time to pregnancy.
- **Mismanagement of Infertility Cases:** Ultimately, diagnostic errors from semen analysis can lead to overall mismanagement of the infertility case. This includes scenarios like treating a couple as “unexplained infertility” (or attributing the issue to the woman) when in fact an undetected male factor exists, or vice versa. A survey of UK laboratories noted that inconsistent adherence to quality standards in semen testing “may have a detrimental effect on result accuracy and consequently lead to patient misdiagnosis and mismanagement”^16^ In other words, unreliable semen analyses can cause clinicians to pursue the wrong course—either over-treatment, under-treatment, or focusing on the wrong partner—jeopardizing the couple’s chance of conception. Given that therapies for male infertility (hormonal treatments, surgeries, ART, etc.) often carry significant side effects and stress, a misdiagnosis places patients at risk of unneeded side effects and emotional turmoil without benefit ^9^.

### Variability in Sperm Distribution

The 2021 WHO laboratory manual (6th edition) underscores semen analysis as the cornerstone of male fertility assessment^17^. Although it mandates standardized protocols to ensure result comparability, current methodologies often fail to meet rigorous statistical standards due to inherent technological constraints. Manual microscopy can require up to 45 minutes per sample, with studies reporting inter-technician variability in the range of 20– 30%.^9,18^

Semen samples, even after homogenization, do not exhibit perfectly uniform distributions across the slide or microchamber. Instead, variations in sperm density occur due to factors such as differential glands of fluid origin, fluid dynamics, sperm motility patterns, and sample preparation inconsistencies. Additionally, spatial clustering effects introduce additional variability into sperm concentration measurements.^19^

To ensure collection of sufficient statistical information, the WHO guidelines for sperm analysis emphasize the importance of counting a sufficient number of sperm cells to achieve reliable measurements. According to the WHO, sperm concentration should be assessed by analyzing at least 200 spermatozoa in a properly prepared sample, and motility should be evaluated based on at least 400 spermatozoa.^17^

To counteract non-uniform sperm cell distribution across the sample and ensure accuracy and repeatability, the WHO recommends performing replicate aliquots analysis to reduce the risk of non-representative sampling. The guidelines also stress the necessity of random field selection to avoid localized sperm clustering effects, which can distort concentration and motility estimates.^20^ For extremely low sperm concentrations such as post-vasectomy semen analysis, the WHO advises using enhanced sensitivity methods requiring observation of as much sample volume as possible.

While the WHO guidelines establish statistically rigorous sperm analysis, the practical implementation with existing analysis methods necessitates the examination of multiple sample areas and FOVs per patient. Therefore, strict adherence to the WHO guidelines extends measurement durations, subsequently elevating analysis costs and diminishing test accessibility..

### Expanded Field of View

To eliminate the need of multi-FOV analyses, we propose illumicell AI’s proprietary LuceDX—an advanced imaging system that captures a significantly expanded FOV of approximately 3×4.2 mm, roughly 13 times greater than the standard 1×1 mm. The larger imaging area preserves resolution comparable to standard CASA systems yet dramatically boosts statistical reliability, as parameters estimations are based on analysis of a significantly bigger number of sperm cells compared to standard methods. By encompassing more of the sample in a single frame, LuceDX mitigates non-uniform distribution biases and clustering effects, consequently reducing measurement error.

These advantages are particularly relevant for oligozoospermic and cryptozoospermic analyses, where sperm counts can be extremely low and erroneous readings may impact clinical care. Standard-FOV CASA can be prone to false negatives or highly variable counts in such diluted samples. By virtue of its expanded coverage, LuceDX captures a broader sample area, offering more reliable detection of sparse sperm and fewer false negatives.

Overall, an expanded-FOV system—combined with robust image analysis—addresses the documented, technical shortcomings of both manual and CASA-based semen assessments. By mitigating variability, misdiagnosis risk, and unnecessary cost, it stands to meaningfully improve the clinical management of male infertility.

## Materials and Methods

### Participant Recruitment, Consent, and Anonymity

Participants were referred by their physicians to the Fertas Laboratory for standard semen analyses. Prior to their scheduled appointments, each potential participant received an informed consent form and was given at least seven days to review and consider the document. Participation in the study was voluntary, and patients could withdraw consent at any time without affecting their medical care.

Upon arriving at the Fertas laboratory, patients were asked to provide their informed consent. If a patient provided consent, only the fraction of the semen sample not needed for routine diagnostic tests (typically 10 µL) was allocated for the investigational analysis. In cases where the entire sample was required for the prescribed medical analyses, the study did not use any material. The samples never left the laboratory, ensuring the continuity and the security of the chain of custody. The described participant recruitment flowchart is depicted in Fig. 1.

**Figure 1.**
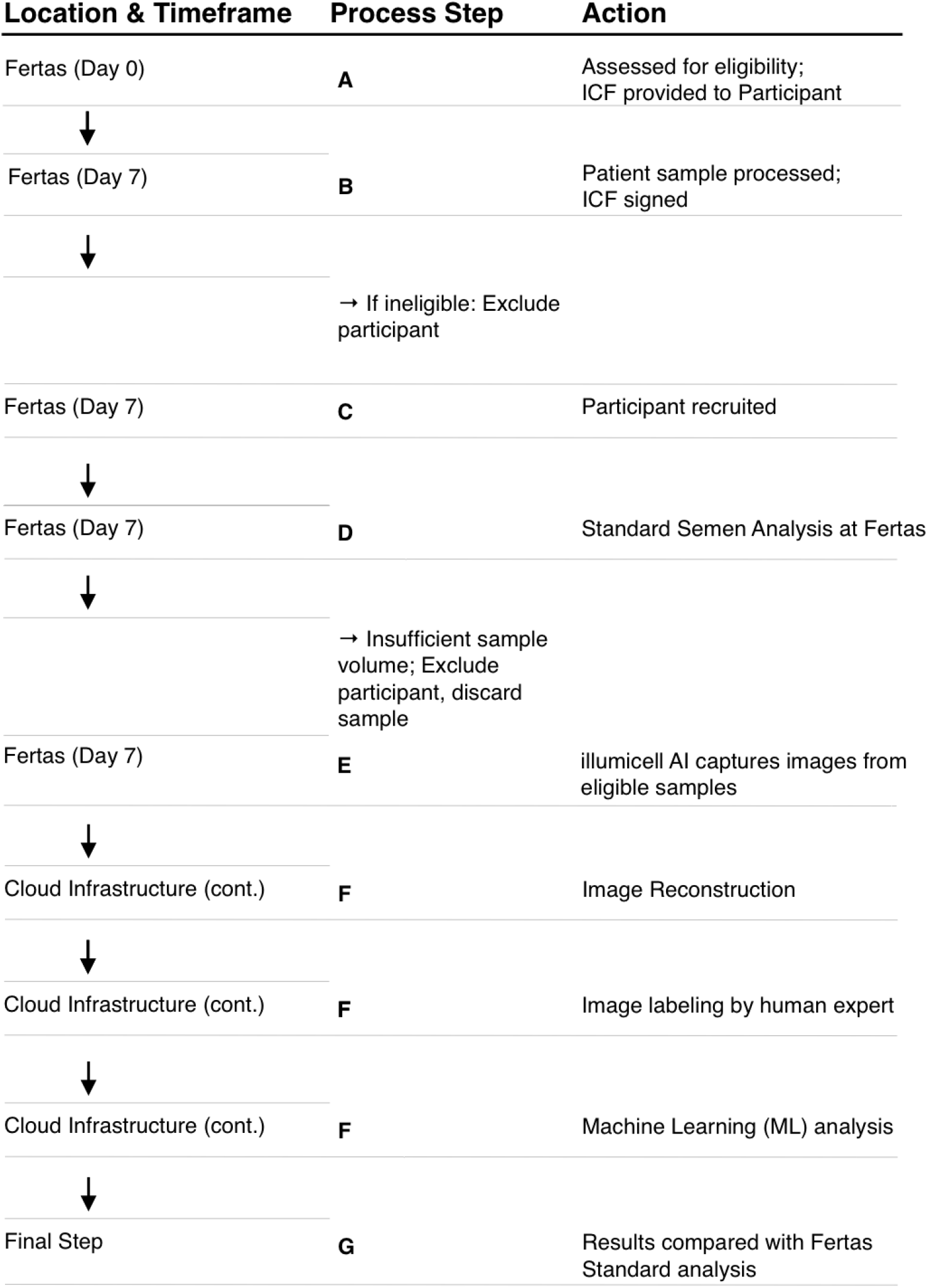
Participants recruitment flowchart. (A) Participants are assessed for eligibility on day 0 and receive the informed consent form. (B) Patient samples are processed and ineligible cases are excluded. (C) The formal recruitment of eligible participants on day +7, followed by (D) samples assesment by Fertas Laboratory. (E) If the sample volume is insufficient, participants are deemed ineligible or the specimen is discarded; otherwise, illumicell AI captures images for (F), a cloud-based process involving image reconstruction, human expert labeling, and continuous machine-learning analysis. (G) Comparison of extracted results with the Fertas assessments.

To protect participant privacy, each patient who consented was assigned a unique random identification number. This coded identifier was used on all research materials and data, ensuring that no personally identifying information was linked to the samples or the analysis. The consent documentation, which contained participant names and signatures, was stored separately at the Fertas laboratory and was not directly associated with the study results. Furthermore, Illumicell AI received only anonymized results of the assessments conducted at Fertas, ensuring that no personally identifiable information was shared. Only the designated site director retained the means to re-identify any data if necessary.

### Sample preparation and homogenization

Semen samples were prepared following the WHO 6th guidelines for semen analysis at Fertas Laboratory. Upon collection, the semen sample was allowed to liquefy at 37°C for 30-60 minutes before analysis. After liquefaction, the total volume of the ejaculate was measured, and the sample underwent thorough mixing using pipetting to achieve homogeneity.

### Concentration and Motility Measurement Protocols

A laboratory technician examined a small portion of the native undiluted sample under a benchtop phase-contrast microscope to perform an initial qualitative assessment. At this stage, the technician measures pH using a test strip and determines the appropriate dilution factor in accordance with World Health Organization (WHO) guidelines and personal expertise.

An aliquot of the sample was subsequently diluted to a concentration of approximately 20-40 million cells/mL and inserted into a 20 μm depth microfluidic chamber (SC20-01C Leja Standard Count 20-micron, 2-chamber slide) for further analyses with the CASA system and LuceDX. Undiluted samples were used for sample presenting lower concentrations.

#### CASA System

(Standard FOV 1×1 mm, Baseline Method): A diluted semen sample was inserted into a Leja microfluidic chamber and analyzed by the CASA system coupled to an optical microscope (10× objective, phase-contrast mode) to measure motility and concentration within a 1×1 mm field of view. The CASA system automatically counted the number of sperm cells per FOV and assessed their trajectories using proprietary algorithms. The technician entered the chosen dilution factor into the system to extract the correct values of concentration and motility and assessed the validity of the automatically generated results. At least 2 FOV were analysed separately per sample. More FOV were used for low semen concentrations. This entire process, from sample insertion to final result recording for concentration and motility, typically requires 5–10 minutes per sample. Once the results were recorded and the Fertas technician ensured that the sample was no longer needed for clinical evaluation of the patient, the sample was coded (anonymized) and transferred to illumicell AI’s technician for further evaluation.

#### LuceDX

(Expanded FOV 3×4.2 mm, Studied Method): Analyses with LuceDX were carried out with a 5–10 minute time delay compared to evaluation with the CASA system. A coded sample was placed in the measurement chamber, and the instrument captured a sequence of full-FOV frames, which were further utilized for concentration and motility analysis. To ensure proper data collection, each sample underwent between two and four measurements. Subsequently, the captured data was uploaded to a secure cloud server in Switzerland for further processing and data analysis. After data acquisition, the sample was discarded and destroyed according to the laboratory’s standard protocols.

### Post-Vasectomy Sample Analysis

To address the extremely low sperm concentration problem encountered in post-vasectomy semen analysis, rendering standard CASA systems unreliable, the Fertas Laboratory implemented a modified protocol adhering to WHO guidelines. This protocol is based on manual analysis and was necessary to accurately assess samples with such low sperm counts. The post-vasectomy sample was allowed to liquefy for 30 – 60 minutes, after which the total ejaculate volume was recorded. A 10 µL aliquot of the native sample was then placed on a standard 76×26 mm microscope slide and covered with a glass coverslip.

#### Manual Analysis

Using a benchtop phase-contrast microscope with a 40× objective, a trained laboratory technician scanned the entire slide to locate any remaining spermatozoa and assess their motility. This process was performed twice, each time with a new 10 µL aliquot on a separate slide. The entire process typically requires up to 20 minutes per sample. The technician’s expertise was critical in detecting and evaluating any residual spermatozoa. Once the results were recorded, the sample was coded (anonymized) and transferred to illumicell AI’s technician for further evaluation.

#### LuceDX

For the LuceDX analysis of post-vasectomy samples, the measurement methodology did not differ from a standard SA. The coded specimen was placed into the measurement chamber, and the system captured a sequence of high-resolution images, typically with two to four measurement sequences per sample. The resulting data was uploaded to the secure Swiss cloud service for processing. Once the measurements were completed, the sample was discarded and degraded according to the standard protocols of our laboratory.

### Collected Data

The data in Table 1 represents the measured and calculated parameters by Fertas Laboratory, as part of an ongoing sample acquisition campaign, with each “N” corresponding to a unique patient. Concentration and motility analyses were performed by Fertas technicians with aid of QualiSperm CASA system.

**Table 1.** Overview of the key SA parameters measured by Fertas Laboratory for the samples collected from recruited participants. The mean value, standard deviation (SD) and range of values are presented for all 44 participants (Total) along with subsets of 21 participants with normal SA parameters (Normal) and 23 participants with abnormal concentration or motility readings (Abnormal).

| Parameter | Total (N=44) |  |  | Normal* (N=21) |  |  | Abnormal* (N=23) |  |  |
| --- | --- | --- | --- | --- | --- | --- | --- | --- | --- |
|  | Mean ± SD | Median | Range (min–max) | Mean ± SD | Median | Range (min–max) | Mean ± SD | Median | Range (min–max) |
| Concentration (×10<sup>6</sup>/mL) | 61.5 ± 70.1 | 51.7 | 1.5–422.4 | 97.5 ± 83.7 | 75.4 | 27.9–422.4 | 28.5 ± 29.3 | 9.6 | 1.5–90.8 |
| Total Motility (%) | 40.7 ± 19.6 | 40.0 | 0.0–74.0 | 56.2 ± 11.2 | 56.1 | 40.5–74.0 | 26.5 ± 14.1 | 26.3 | 0.0–66.1 |
| Progressive Motility (%) | 36.5 ± 18.1 | 34.4 | 0.0–70.7 | 50.3 ± 11.5 | 49.3 | 32.5–70.7 | 23.8 ± 12.9 | 24.3 | 0.0–58.9 |

*\* Abnormal SA readings were defined as concentrations below 16 million cells/mL or total motility below 40%, as outlined in the WHO 6 guidelines.*

In this cohort, 52% of the samples present either concentrations below 16 million cells/mL or total motility below 40%, rendering them abnormal by falling under the WHO 6 reference values. The remaining 48% of patients exhibited concentration greater than 16 million cells/mL and total motility greater than 40% which aligns with the normal thresholds set by the WHO 6 guidelines. These samples were deemed as “normal”. It is important to note that this designation does not constitute a full diagnostic judgment, as additional factors such as pH levels, morphology, and other clinical considerations may be necessary to fully assess each individual reproductive health. Nevertheless, the diversity of data acquired enabled us to test our system. These parameters represent not only ideal “best-case” scenarios but also more challenging samples that require careful analysis.

### Assessment of FOV Effects in Sperm Analysis

Sperm concentration is one of the most important parameters in SA and male fertility evaluation, serving as a major indicator for infertility treatment. With the WHO establishing a lower reference limit of 16 million cells/mL for normal SA^17^, existing CASA systems often struggle to provide accurate counts at these concentrations, potentially leading to misdiagnosis and inappropriate treatment decisions. Moreover, precise concentration analysis is particularly crucial in post-vasectomy SA, where concentrations below 0.1 million cells/mL must be reliably detected to confirm successful vasectomy. The absence of current technical solutions to provide reliable assessment of sperm concentration pushes clinicians to rely on labor-intensive manual analyses.

In this section, we discuss the role of the FOV on the precision of semen analysis across a spectrum of sperm concentrations. This is to address the critical challenge of mitigating the medical and labor costs in semen analysis. We took a statistical treatment approach and demonstrated an example of the utility of LuceDX on how a system with an enhanced FOV can contribute to increased precision of concentration assessment.

### Effect of FOV on Measurement Precision

In the context of sperm analysis, concentration is typically defined as the number of sperm cells per ejaculate volume and is measured in millions of sperm per milliliter (M/mL). However, a single sperm concentration measurement using a microscopic technique involves counting sperm cells within a given FOV. Therefore, the experimentally measured concentration of sperm cells C_M_ is defined as

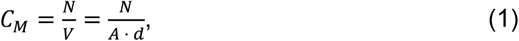

where *N* is the number of observed sperm cells, and *V* is the sample volume, which can be expressed as a product of *A*, the area of the FOV, and *d*, the thickness of the sample layer.

To quantify the concentration measurement precision, we use the Relative Standard Error (RSE). RSE is a crucial metric in assessing the precision of experimental measurements, as it demonstrates the ratio between the error and the measured value, providing a way to directly compare measurements with different parameters. RSE is defined as the ratio between the standard error of the measured value, *SE(C*_*M*_), and the value itself,

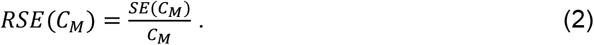

Let us first assume the ideal case in which the sperm cells are randomly distributed across the sample, i.e., their distribution follows Poisson statistics. Then, using the equality between the variance and the mean for the Poisson distribution, *Var*(*N*) = *C* · *V*, where *C* is the true concentration value, the standard error can be expressed as

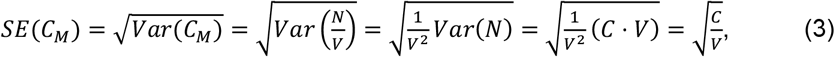

Then, the RSE can be rewritten as

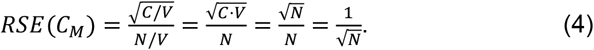

Under the assumption of a random sperm distribution with a given concentration, the number of sperm cells present in the image is directly proportional to the area of the FOV. Consequently, if a baseline FOV contains *N*_*0*_ sperm cells, increasing the FOV by a factor of *k*_*FOV*_ results in a proportional increase in the number of sperm, *N = k*_*FOV*_ *· N*_*0*_. Therefore, the RSE for a system with a larger FOV can be expressed as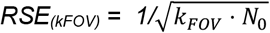.

Lastly, we introduce the RSE reduction factor *ξ*, defined as

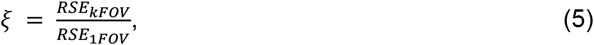

which represents the reduction in the variability of measurement outcomes in systems with different FOVs. After substituting equation (4) into equation (5), the RSE reduction factor becomes

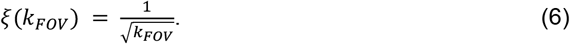

Equation (6) demonstrates that, under the assumption of a random sperm distribution with a given concentration, the reduction in SEM arises solely from the FOV scaling factor *k*_*FOV*_. Practically, this means that a larger FOV significantly enhances the precision and reliability of semen assessments by reducing *ξ*, effectively minimizing the variability in measurement outcomes arising from a limited sampling statistic. Figure 2, which graphically depicts equation (6), illustrates how increasing the FOV systematically decreases the relative measurement error.

**Figure 2.**
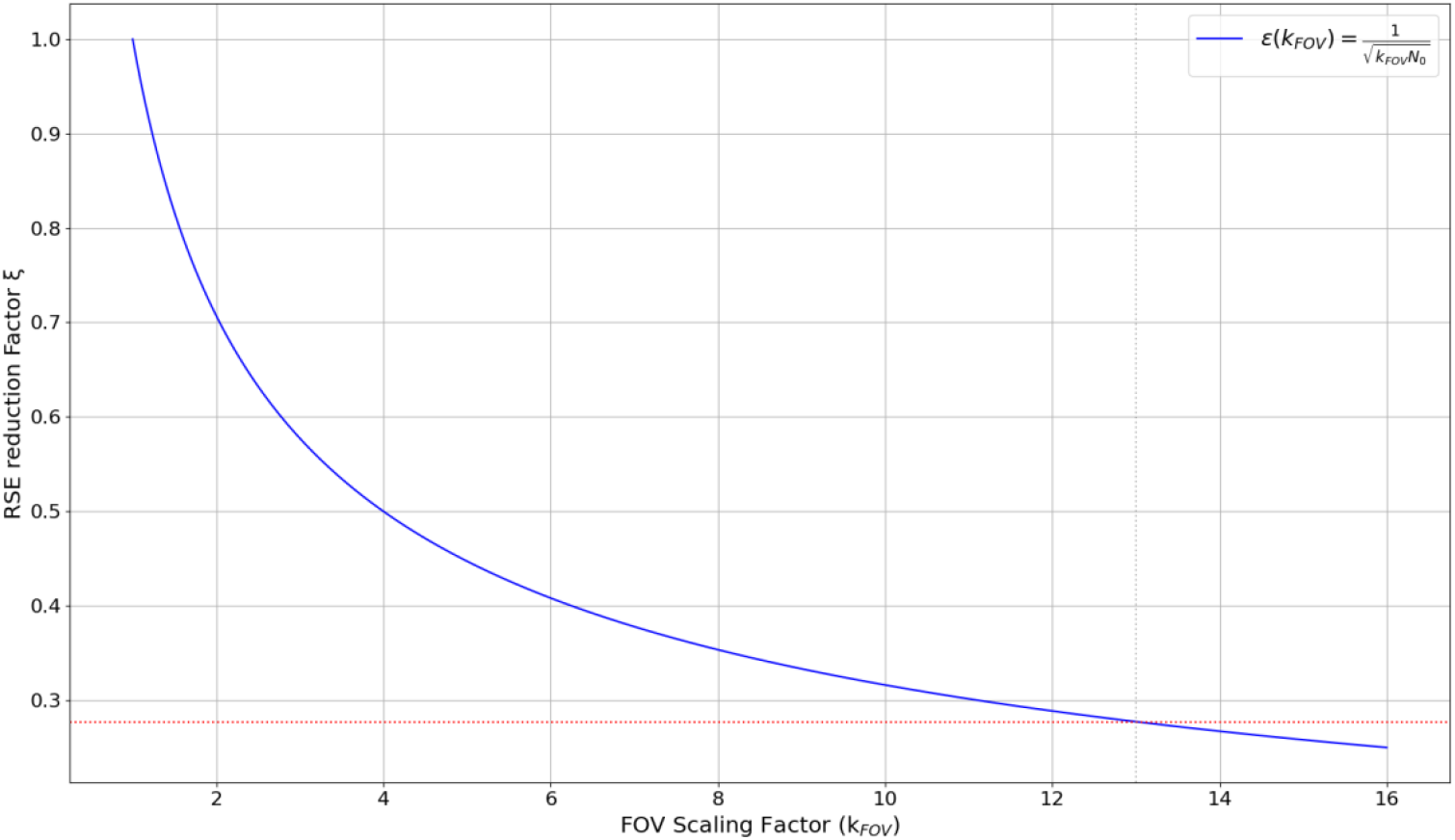
Impact of field of view (FOV) on the relative standard error (RSE) reduction factor ξ. LuceDX enhanced FOV (k_FOV_ = 13) reduces the measurement variability by more than 3 times compared to a standard CASA system (red dashed line).

As LuceDX demonstrates a 13-fold increase in FOV compared to a standard CASA system, the RSE reduction factor *ξ* for this pair becomes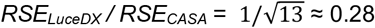. This practically implies that a single image analysis of the same sample performed with LuceDX would yield a measurement precision approximately 3.6 times greater than that obtained from a standard CASA system, directly contributing to a more reliable estimation of sperm concentration. Furthermore, the utilization of a system with an expanded FOV may reduce the need for analyzing multiple sample areas, thereby saving the operator’s time.

### Effect of Concentration on Measurement Precision

In the previous section, we examined the sole influence of FOV on the RSE reduction factor *ξ* under the assumption of uniform sperm distribution for a given patient, fully omitting the influence of the sperm concentration itself. However, the patient’s condition and the associated sperm concentration play an important role in determining measurement precision.

As the number of sperm cells counted in a given field of view, according to the equation (1), can be expressed as

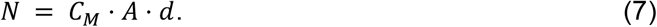

If both concentration and FOV can vary, recalculating the SEM reduction factor *ξ* requires adjusting the number of cells not only by the FOV scaling factor *k*_*FOV*_ but also by a concentration factor, *C*_*F*_, compared to standard parameters, *N*_*0*_ *= C*_*0*_ · *A*_*0*_ · *d*. This results in an increase in the number of observed sperm cells, *N = k*_*FOV*_ *· C*_*F*_ *· N*_*0*_, with an increase in either FOV or sperm concentration. The SEM reduction factor *ξ* then becomes

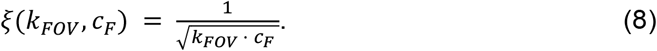

### Low Count and Post-Vasectomy Sperm Analysis

The practical implications of FOV and concentration effects on measurement precision manifest themselves most strongly in SA for patients with lower sperm concentrations. In typical semen samples from fertile or subfertile individuals, sperm concentrations range from 16 to 100 M/mL. Within this range, the change in sperm count precision due to concentration variations is relatively modest, and the RSE reduction factor, corresponding to a 5x *C*_*F*_ variation for a given FOV, does not exceed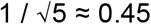 Moreover, to preserve the number of sperm cells per FOV, many clinics implement an additional patient-specific ejaculate dilution, which further minimizes the concentration influence in real measurements.

However, the role of concentration in measurement precision becomes critical when the concentration approaches the lower reference limit of 16 M/mL, and especially in post-vasectomy semen analysis with typical concentration readings below 0.1 M/mL. At these low concentrations, the number of sperm cells captured within a standard FOV becomes exceedingly small, leading to significant variability in counts.

To illustrate this effect, we plot RSE reduction factor *ξ*, defined by equations (5) and (8), as a function of sperm concentration for various FOV scaling factors *k*_*FOV*_ (Figure 3). For a baseline *RSE*_*1FOV*_, we chose the case of an average patient with a standard CASA FOV: the number of observed cells for this case can be expressed as *N*_*0*_ *= C*_*0*_ · *A*_*0*_ · *d*, where *C*_*0*_ = 40 M/mL and *k*_*FOV*_ = 1 (*A*_0_ = 1 × 1 mm^2^).

**Figure 3.**
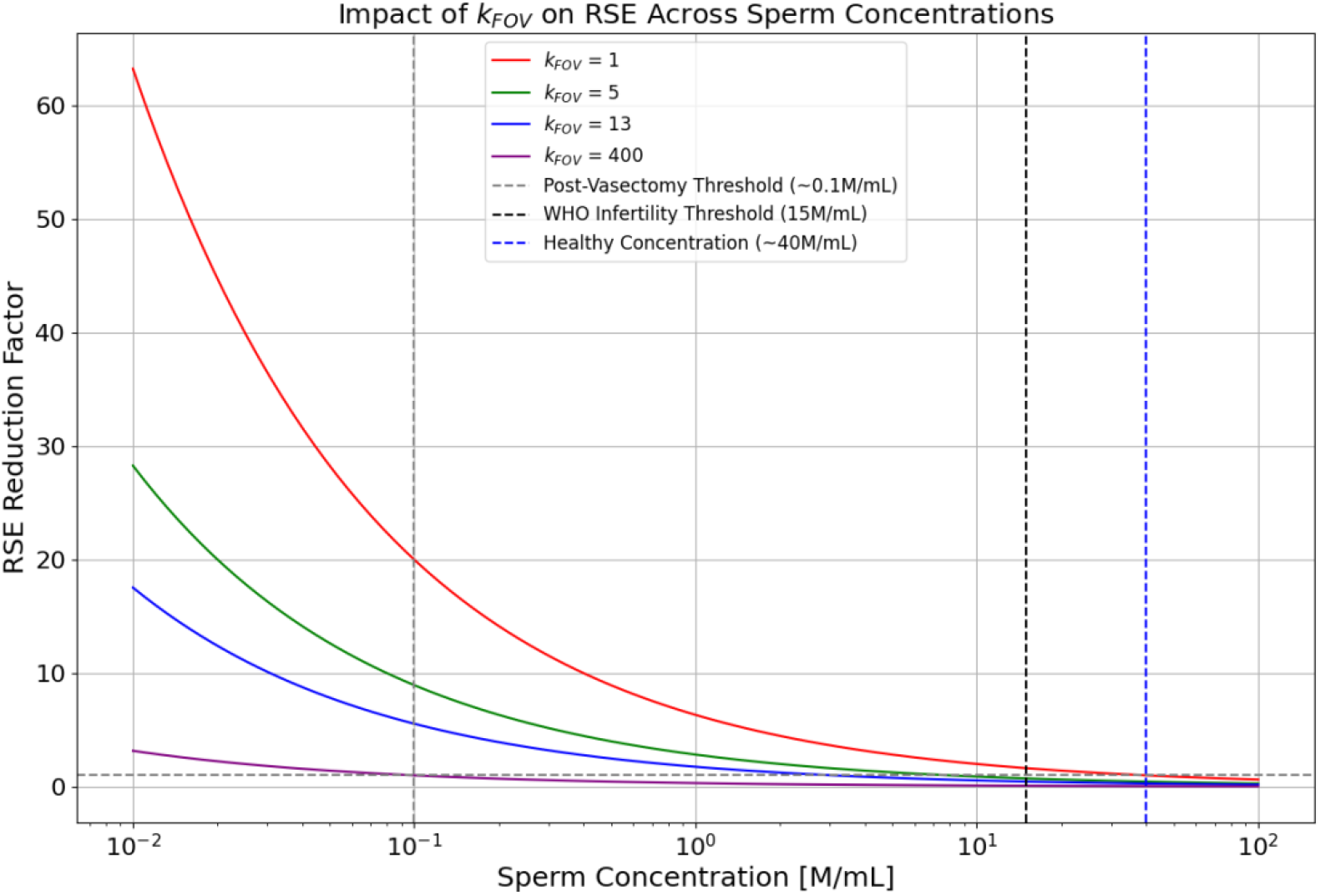
Impact of sperm concentration and FOV scaling factor k_FOV_ on the RSE reduction factor (ξ). Owing to its expanded FOV, LuceDX with the FOV scaling factor k_FOV_ = 13 provides the same measurements precision at concentration level of 3 M/mL as standard CASA systems with the FOV scaling factor k_FOV_ = 1 at 40 M/mL. Vertical dashed lines indicate key concentration levels: 0.1 M/mL (light grey), 16 M/mL (dark grey), and 40 M/mL (blue), representing the post-vasectomy threshold, lower reference limit, and average normal concentration, respectively. The horizontal grey dashed line (ξ = 1) represents the baseline RSE. LuceDX, with its 13x FOV scaling factor, demonstrates enhanced precision across a range of concentrations, particularly at the critical 0.1 M/mL level.

= 40 M/mL and *k*_*FOV*_ = 1 (*A*_*0*_ = 1 × 1 mm^2^).

Figure 3 can be interpreted in the following way: whenever the RSE reduction factor is below the *ξ = 1* line (horizontal grey dashed line), the precision of the measurement is improved compared to the baseline precision; when the RSE reduction factor is above the *ξ = 1* line, the precision is declined; for the baseline case *k*_*FOV*_ = 1 and *C*_*0*_ = 40 M/mL the RSE reduction factor equals to 1.

The plot highlights three key concentration levels, marked by vertical dashed lines. The 16 M/mL (dark grey dashed line) and 40 M/mL (blue dashed line) concentration levels represent the lower reference limit and average normal concentration readings, respectively. It is evident that even a 5x FOV increase can elevate the precision of threshold measurements to levels comparable to those achieved with a standard FOV. With a 13x FOV increase, as demonstrated by LuceDX, the precision of concentration measurements remains above the standard precision level down to a 3 M/mL concentration. Practically, this means that semen analyses performed with the LuceDX system are significantly less susceptible to the decrease in precision observed with standard CASA systems, thereby reducing the risk of misdiagnosing patients.

The final highlighted concentration level, 0.1 M/mL (light grey dashed line), represents the post-vasectomy concentration threshold. As the plot indicates, achieving the precision of a standard analysis at this low concentration would necessitate a 400-fold increase in FOV, a feat currently beyond the capabilities of existing technologies. However, improved precision at this concentration is of paramount importance, as post-vasectomy analysis currently presents one of the most challenging scenarios for SA. Notably, the LuceDX system, with its 13-fold increase in FOV, significantly enhances precision at this critical concentration level, bringing it to the level achievable by CASA systems at 1 M/mL levels, thereby addressing a key limitation in current diagnostic approaches

### Assessment of Clustering Effects in Sperm Analysis

So far, we have assumed perfectly random distributions in the observed sperm samples. However, even with ideal homogenization of the sample, the assumption of a Poisson distribution does not hold in practice. Instead, in real samples, local density fluctuations arise due to biological and physical effects. Motile sperm accumulate in specific regions due to hydrodynamic forces and chemotaxis, creating localized areas of higher concentration.

To quantify the impact of clustering on concentration estimates, we introduce the variance-to-mean ratio (*VMR*), a measure of the degree of clustering (overdispersion) in the sperm cell distribution, defined as

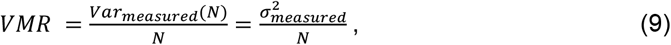

where 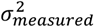 is the measured variability of the sperm counts across multiple FOVs, reflecting how much the counts deviate from their average. A value of *VMR* > 1 indicates clustering, meaning sperm tend to accumulate in localized regions, *VMR* = 1 suggests a perfectly random Poisson distribution, and *VMR* < 1 reveals a more dispersed, regular sperm distribution.

To account for the clustering effect, we re-evaluate the RSE defined by equation (2), omitting the assumption of equality between the variance and the mean used in equation (3), which held for the Poisson sperm distribution, *Var*(*N*) = *C* · *V*, and adjusting it to the VMR factor, *Var*(*N*) = *VMR* · *C* · *V*. Then, the RSE becomes

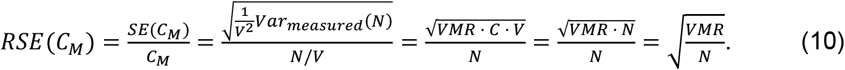

The relationship between the VMR and the FOV depends on multiple factors, such as the cluster size and the exact spatial distribution, and lies outside of the scope of this study. Nevertheless, to illustrate the qualitative impact of the VMR on measurement precision and the corresponding reduction in RSE with increasing FOV, we demonstrate three representative samples from our dataset: Samples 1-3, corresponding to A295, A262, and A898 patients. Figure 4 features heatmaps of sperm distributions for Samples 1-3 (top row), with the full FOV of 3 × 4.2 mm^2^ that reveal distinct spatial patterns. These heatmaps illustrate sperm cell distributions with varying degrees of clustering: moderate, pronounced, and practically absent for Samples 1-3, respectively. For each heatmap, we provide a histogram of sperm counts within a 55 × 55 µm^2^ moving window (bottom row), superimposed with a Poisson distribution fit (red line). These histograms demonstrate how closely the experimentally assessed distribution aligns with the Poisson distribution, with Sample 2 showing the poorest agreement with the Poisson distribution curve.

**Figure 4.**
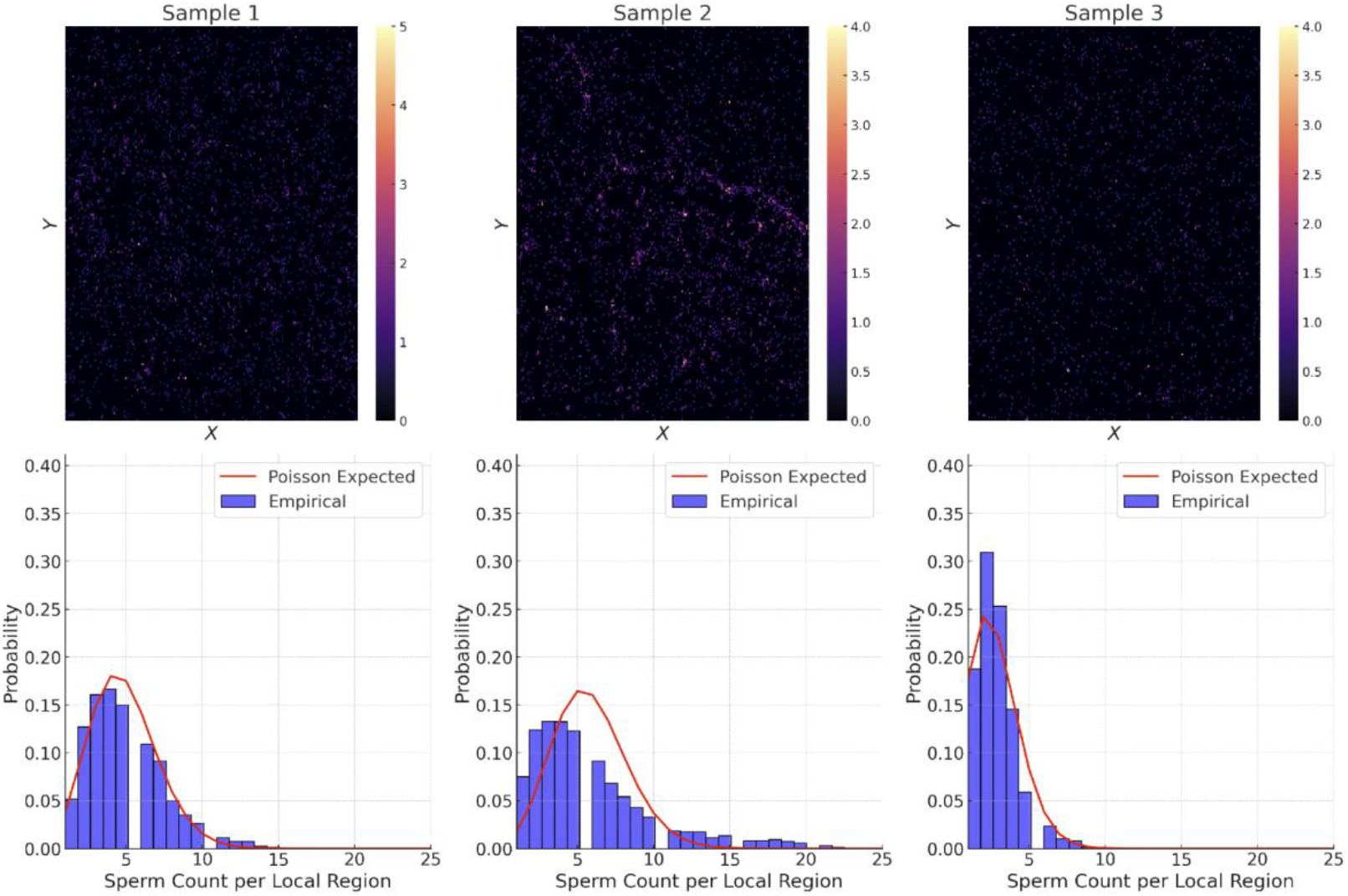
Sperm cell distributions for Samples 1-3 that illustrate a variability of sperm cell spatial distributions present in the collected dataset. Top row: Heatmaps of sperm cell distributions with varying degrees of clustering: moderate (Sample 1), pronounced (Sample 2), and practically absent (Sample 3). Bottom row: Histograms of sperm counts within a 55 × 55 µm^2^ moving window, superimposed with a Poisson distribution fit (red line).

To quantitatively analyze these spatial distributions, we computed the mean number of sperm cells per FOV, N_mean_, the measured standard deviation, σ_measured_, and the VMR for a moving window with a standard FOV of 1 × 1 mm^2^, evaluated at 50 different locations. The summary of these statistical quantities for Samples 1-3 is presented in Table 2.

**Table 2.** Summary of statistical quantities for Samples 1-3, including the mean number of sperm cells per FOV (N_mean_), the measured standard deviation (σ_measured_), and the Variance-to-Mean Ratio (VMR), evaluated at 50 different FOV locations. Calulated VMR parameters serve as the main identifier for the spatial distribution type.

| Sample | $N_{\text{mean}}$ | $\sigma_{\text{measured}}$ | VMR |
| --- | --- | --- | --- |
| Sample 1 | 281 | 21.4 | 1.6 |
| Sample 2 | 295 | 49.8 | 8.4 |
| Sample 3 | 144 | 12.0 | 1.0 |

Sample 1, for which the histogram in Figure 4 closely follows the Poisson fit, exhibits a VMR of 1.6, representing an example of a spatial distribution with mild clustering. Sample 2 deviates strongly from the Poisson model, displaying significant overdispersion with a VMR of 8.4, and exhibiting strong sperm accumulation in localized regions. In contrast, Sample 3 exhibits an almost ideal fit with a VMR of 1, representing an example of a true Poisson distribution. These results demonstrate the variability in sperm spatial distributions and emphasize the need for clustering-aware statistical corrections in sperm concentration estimation.

To illustrate the influence of FOV on measurement precision for different degrees of spatial clustering, we compared concentration estimates using a moving window corresponding to *k*_*FOV*_ *=* 1,5, and 8, with respect to a standard FOV of 1×1 mm^2^, as shown in Figure 5. To mimic real-world variability introduced by operators or lab technicians, each FOV and sample was evaluated at 50 different FOV locations, represented by boxplots. In these boxplots, the interquartile range (IQR)—defined as the range between the 25th and 75th percentiles—captures the middle 50% of the data and serves as a robust visual measure of variability, less sensitive to outliers than standard deviation. For better comparison, concentrations are normalized with respect to the concentration obtained for the full FOV for each sample. The results confirm that FOV size has a significant impact on measurement stability, particularly in highly clustered samples. Although Sample 1and Sample 2 have similar sperm concentrations, their estimation precision differs markedly due to spatial organization. For small FOVs (*k*_*FOV*_*=1*), Sample 1 shows a normalized IQR of approximately 0.76 to 0.88, which tightens to 0.89 to 1.04 at *k*_*FOV*_*=8*, reflecting improved measurement stability. Sample 2, exhibiting stronger clustering, displays a broader IQR at *k*_*FOV*_*=1* (0.66 to 1.03), which remains relatively wide even at *k*_*FOV*_*=8* (0.86 to 1.09), indicating persistent variability despite increasing FOV size. In contrast, Sample 3, with a much higher concentration (also prepared with a larger dilution factor)and exhibiting an almost ideal Poisson distribution, shows narrow IQRs across all scales - from 0.94 to 1.01 at *k*_*FOV*_*=1* to 0.98 to 1.00 at *k*_*FOV*_*=8*. These observations demonstrate that accurately capturing the degree of clustering is more critical to concentration estimate precision than the concentration level itself, especially when using small sampling areas.

**Figure 5.**
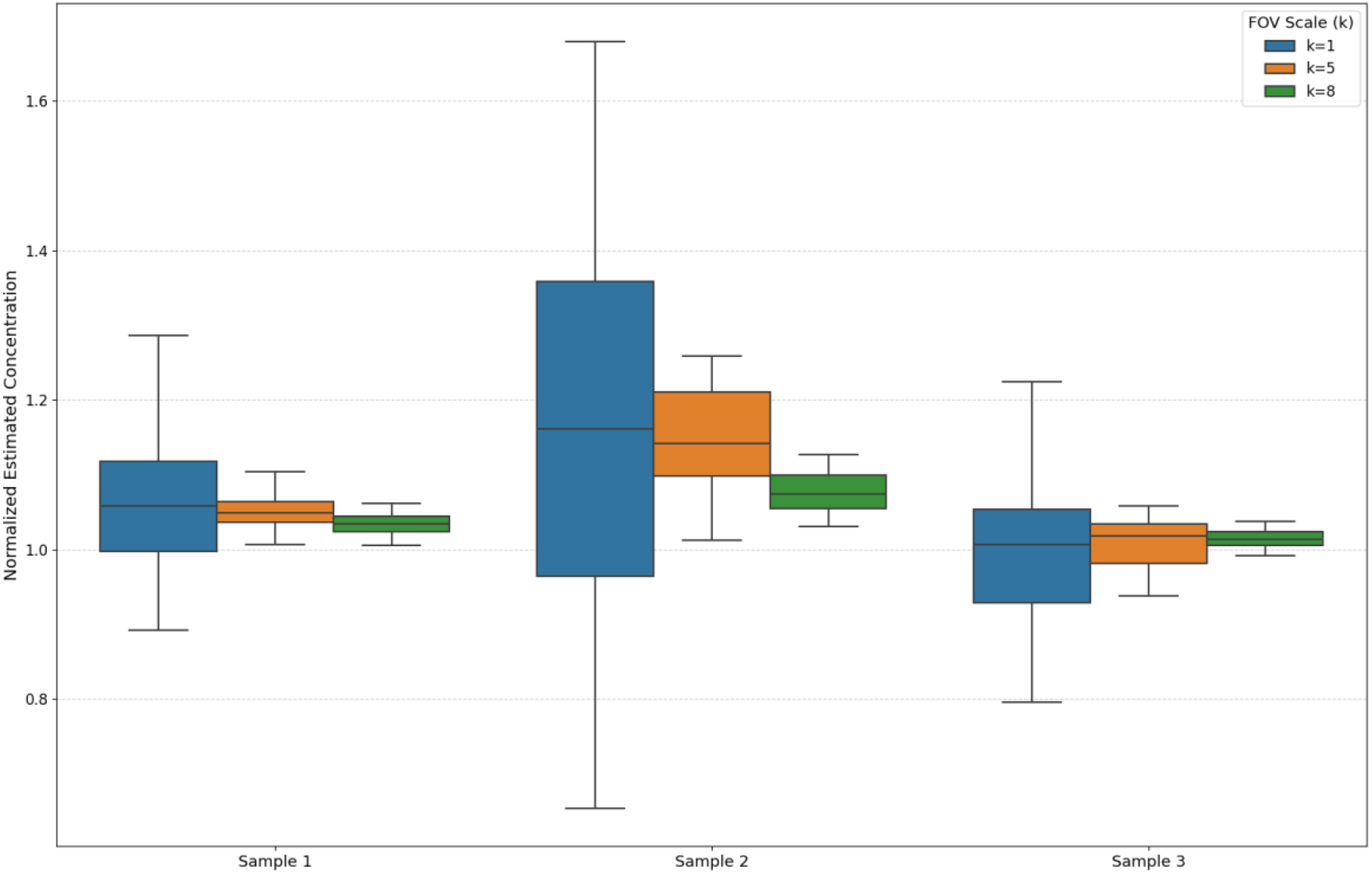
Comparison of concentration estimates for Samples 1-3 demonstrates precision decline for highly clustered samples (Sample 2). Precision decline can be partially counteracted by FOV enhancement, as the spread of concentration estimates decreases with increasing k_FOV_. Comparison of concentration estimates for Samples 1-3 has been done using a moving window corresponding to k_FOV_ = 1, 5, and 8, with a standard FOV of 1 × 1 mm^2^. Boxplots represent variability across 50 different FOV locations. For better comparison concentrations are normalized with respect to the concentration obtained for the full FOV for each sample.

These findings emphasize that the use of smaller FOVs leads to unreliable concentration estimates in clustered samples, whereas expanding the FOV stabilizes measurements by averaging over larger spatial regions. However, even in highly clustered samples, some residual variability remains, indicating that clustering-aware statistical corrections may further improve measurement reliability.

## Discussion

The current study underscores the putative clinical and economic consequences of inaccurate semen analysis, and subsequent overtreatment or undertreatment in infertility management^14,21^. Manual microscopy—long considered the gold standard—remains vulnerable to operator-dependent errors and extended sample processing times^4^. Meanwhile, CASA systems, although faster and more standardized, still exhibit limited accuracy gains over manual analysis, especially in abnormal or low-concentration semen samples^9^. Consequently, a limited field of view emerges as a critical factor that inherently constrains statistical robustness, regardless of the method used.

Our findings indicate that enlarging the FOV from approximately 1×1 mm (typical of standard CASA) to around 3×4.2 mm (as in LuceDX) reduces measurement variability by a factor of about 3.6. This improvement aligns with sampling theory (RSE ∝ 1/√N): capturing more sperm in each frame diminishes the relative standard error. Notably, this effect persists even when accounting for spatial clustering, which arises from factors such as motility-driven micro-aggregates or uneven slide loading^19^. By covering a larger portion of the slide, the system mitigates local density fluctuations—an advantage especially pertinent in extremely low-concentration contexts like post-vasectomy checks and oligospermic versus azoospermic patients, where missing even a few sperm may alter clinical decisions.

Clinically, adopting an expanded-FOV approach may reduce the risk of misdiagnosis and in some cases costly downstream interventions. Inaccurate semen analysis can prompt unnecessary IVF or ICSI cycles, surgeries (e.g., varicocelectomy), or incorrect female-focused treatments if a male factor problem goes undetected^9,15^. The advantage of this approach is especially pronounced in post-vasectomy scenarios, where small-FOV devices tend to yield higher variability or false negatives. With an expanded FOV that captures more representative samples, LuceDX may offer a more reliable detection of residual sperm.

Nevertheless, our findings should be considered preliminary until validated by larger, multicenter studies that include diverse patient groups (e.g., severe oligozoospermia, asthenozoospermia). Although our theoretical models and pilot data support the advantage of an expanded FOV, real-world clustering factors can be highly variable. Comprehensive AI-driven segmentation or adaptive sampling strategies may further mitigate clustering-related errors and improve throughput. Additionally, while we highlight potential cost efficiency of adopting an expanded-FOV system, formal health-economic analyses are warranted to quantify its broader impact on infertility care. Such studies might compare initial capital investment in advanced imaging systems in relation to optimized fertility, shorter time to appropriate interventions.

In summary, our results suggest that an enlarged FOV, exemplified by LuceDX’s ∼13-fold increase in sample coverage compared to traditional CASA, helps overcome recognized shortcomings of both manual and automated analyses. By improving measurement precision, reducing the need for multiple fields, and mitigating clustering effects, an expanded-FOV system has the potential to enhance diagnostic reliability and clinical outcomes in male infertility assessment. Future research should aim to confirm these benefits across larger cohorts, explore integration with advanced image processing, and assess cost-effectiveness in routine clinical practice.

## Data Availability

The data used in the analyses of the present study are available from the corresponding author upon reasonable request.

## Ethical Approval Statement

This study received ethical approval from the Commission cantonale (VD) d’éthique de la recherche sur l’être humain (CER-VD) in Lausanne, Switzerland (Project ID: 2023–02302), titled “Assessment of a Machine Learning Assisted Sperm Analysis System (MASA) for Motility, Concentration, and Morphology.” The authorization is valid for the duration of the study, up to a maximum of 5 years from the date of approval. All procedures involving human participants were conducted in strict accordance with relevant Swiss guidelines and regulations.

